# Intersectoral collaboration and community voice in a marginalized neighbourhood: A longitudinal social network analysis

**DOI:** 10.64898/2026.04.10.26350579

**Authors:** Roos van Lammeren, Jelmer Schalk, Suzan van der Pas, Jet Bussemaker

**Affiliations:** Department of Public Health and Primary Care, Health Campus the Hague, Leiden University Medical Centre, 2511 DP The Hague, The Netherlands; Faculty of Social Work and Applied Psychology, Leiden University of Applied Sciences, 2333 CK Leiden, The Netherlands; Institute of Public Administration, Leiden University, 2501 EE Den Haag, The Netherlands

## Abstract

In this article we argue that intersectoral collaboration is ultimately manifested at the neighbourhood level, where professionals from diverse sectors engage in a joint network to improve population health outcomes. To strengthen intersectoral collaboration in neighbourhoods with low SES, it is crucial to include the community voice, representing diverse citizens who must be heard and engaged in decision-making processes. This study aims to contribute to the literature of intersectoral collaboration by exploring how networks emerge and evolve over time. We focus on the development of the roles of citizens in the professional network and diverse sectoral involvement within a local network of the team called *The Connectors*, in a neighbourhood with low socioeconomic scores (SESs). Methodologically, we use a combination of social network analysis (SNA) and action research. Results show that the network expanded significantly over time, both in terms of the number of actors and the diversity of sectors involved. At both measurement points T1 and T2, the majority of collaborations occurred across sectoral boundaries. By the second measurement, the proportion of intersectoral relationships had increased. This indicates that as the network expanded, new collaborations were not confined to existing sectoral clusters but increasingly bridged different sectors. The dual role that citizens have taken on during the development of the network, serving both as community voice representatives and professionals, can be empowering, offering pathways for personal growth and career advancement. However, it also introduces complexity, as these individuals may experience tensions between personal commitments and professional responsibilities. To enable network development, policies should allow room beyond standard protocols and organizational silos, as well as provide sufficient time for relationships and structures to mature. Although network building is a gradual and complex process, once established, these networks can play a pivotal role in delivering integrated and responsive care.

## Introduction

Collaboration between professionals of diverse sectors is widely advocated to improve population health outcomes (1). Health outcomes are determined by a wide range of factors extending beyond the health sector. Especially in neighbourhoods with low socioeconomic scores (SES), non-medical factors such as income, housing situation, and food insecurity negatively influence health outcomes of the individuals living in those neighbourhoods (2). Therefore, intersectoral collaboration and solutions are needed in such neighbourhoods to improve health outcomes.

In this article we argue that intersectoral collaboration is ultimately manifested at the neighbourhood level, where frontline professionals from diverse sectors engage in a joint network to improve service delivery. Frontline professionals are public-sector workers, including health and social care practitioners, who engage face-to-face with clients (3, 4). The joint network does not emerge naturally, as it is often hindered by institutional boundaries, differing professional logics, and sector-specific priorities (5). Research on local intersectoral collaborations shows that low SES populations are often serviced by a multitude of professionals from diverse sectors, but without those professionals interacting with each other in a substantive way (6). The absence of collaboration between professionals of different sectors limits the effectiveness of service delivery (6). It is therefore important to identify and strengthen collaborative structures in local intersectoral networks of professionals (7).

To strengthen intersectoral collaboration in neighbourhoods with low SES, it is crucial to include the community voice, representing diverse citizens who must be heard and engaged in decision-making processes (8). The inclusion of the community voice can be achieved by collaboration with citizens, because citizens have tacit knowledge of the context and the needs of the neighbourhood they live in (9). Citizens are often referred to as ‘experts by experience’ (10). Collaboration of professionals from a variety of sectors with citizens thus helps to align local client needs and improve care (9, 10). Building on the established understanding that effective collaboration contributes to better outcomes, our focus is on examining how such collaboration emerges and develops over time. In doing so, we aim to advance knowledge on how professionals and citizens can work together more effectively in practice.

Previous network research in health has examined what professional networks look like at a certain point in time, but few longitudinal studies aim to understand the development and sustainability of an intersectoral network from its conception (7, 11, 12). The use of network mapping at multiple moments in time may yield actionable insights that are useful in the context of intersectoral collaborations (13, 14). Networks are dynamic structures with relationships created and dissolved over time (15). Consequently, a single measurement may provide a distorted picture of the actual network dynamics. Multiple measurement points, by contrast, can reduce bias in the interpretation of relational patterns and structural changes, because the fluctuations and temporal variation are captured more accurately (7, 16). Furthermore, by using multiple measurements, changes in relationship patterns and growth processes can be mapped out, which can help policy makers develop targeted interventions to improve collaboration.

This study aims to contribute to the literature on intersectoral collaboration that include community voices. We aim to explore how networks emerge and evolve over time. We focus on the development of the roles of citizens in the network and diverse sectoral involvement within a local network in a neighbourhood with low socioeconomic scores. Particular attention is paid to the dynamics between professionals and citizens who represent the community voice, and how their collaboration deepens and transforms as the network matures. This is studied in a local intersectoral network that aims to identify client needs and effective services regarding (mental) health problems, as an exemplary initiative for intersectoral collaboration including the community voice. We have monitored the network since the initial phase in 2022 up until 2025.

## Method

### Study setting

This study is conducted in Moerwijk, a low-SES neighbourhood in The Hague, which is the third-largest city in the Netherlands (17, 18). The research focusses on the network of “The Connectors” (in Dutch: “De Verbinders”), an interdisciplinary team working in the neighbourhood to provide accessible mental health support in the neighbourhood. The team provides this accessible care by building a network of relevant partners whose expertise matches the needs of the citizens. At the start of the project, the team consisted of five members. The two team leaders are a community builder of the neighbourhood (from the municipality’s social sector) and a health professional in the service of the municipality. The other team members include a psychotherapist, a social worker and a citizen representative serving as the community voice from the neighbourhood. The Connectors started in September 2022. The team aims to be easily accessible for citizens through their active presence in the neighbourhood, for instance in community centres and other meeting points for citizens. But the team is also easily accessible by bringing service organizations together on a case-by-case basis. In this, they fulfil a broker role. The team does not rely on fixed protocols, leaving room for tailor-made solutions for service delivery. Moreover, engagement and support do not require prior sign-up or registration. An appropriate short– and long-term solution is assessed for each citizen requiring care.

The requests The Connectors receive from citizens range from issues facing a group of citizens collectively, such as grief counselling after the death of a community centre volunteer, to individual mental health needs. Questions often contain an underlying problem that requires multiple sectors to resolve. These include institutions from the housing sector to the youth and education sectors and for example financial institutions. To address citizens’ needs, The Connectors work with professionals from these various sectors as well as citizens in the neighbourhood. The team has overarching evidence informed aim to bring the network of citizens and professionals closer together in a way that allows them to complement one another when providing support in the neighbourhood. Therefore, the network of The Connectors is typical of intersectoral networks where professionals from different sectors and various citizens collaborate to achieve joint health outcomes (19). In this study we define a frontline professional as an individual who has a formal appointment in a legally autonomous organization, such as municipality, a social enterprise or a mental health institution. A representative of the community voice is a citizen with an active role in the neighbourhood, but who is formally employed by an organization.

### Study design

We study the network of The Connectors using a social network analytical approach (SNA). SNA is a method that measures and analyses (social) relationships within a network (13, 20). It is used to graphically map actors in a network and the relations between those actors (12). In addition to the visualization of the network, SNA allows for the measurement, description, and interpretation of patterns of relationships in the network and, consequently, understanding complex interactions (13, 21). SNA is particularly well-suited to capture the emergence, strengthening, or weakening of (cross-sectoral) relationships over time, making it a valuable tool for analysing the evolving structure of intersectoral networks (13).

The network data were collected in the context of a broader action research methodology. Both the researchers and the team The Connectors work together in this design and focus on positive and goal-oriented change in collaboration and reflection (22). Action research is a joint and iterative process and executes a cycle of activities, including problem diagnosis, action intervention and reflective learning (23).

#### Network data collection

Network data were collected in two phases (from December 2023 until April 2024, and from Augustus 2024 until March 2025) to capture the structure and composition of the network of professionals and citizens involved. Each phase of network data collection consisted of several steps (see Figure 1). First, individual interviews with the team members of The Connectors took place in rounds 1A and 2A. After that, we analysed these data and made decisions concerning the inclusion of individuals for the round of interviews 1B and 2B via a snowball method. This was followed by the overall network analysis of phase 1 and 2, which we then used for the network reflection sessions 1C and 2C with The Connectors. This procedure resulted in two distinct networks at T1 and T2.

**Figure 1.**
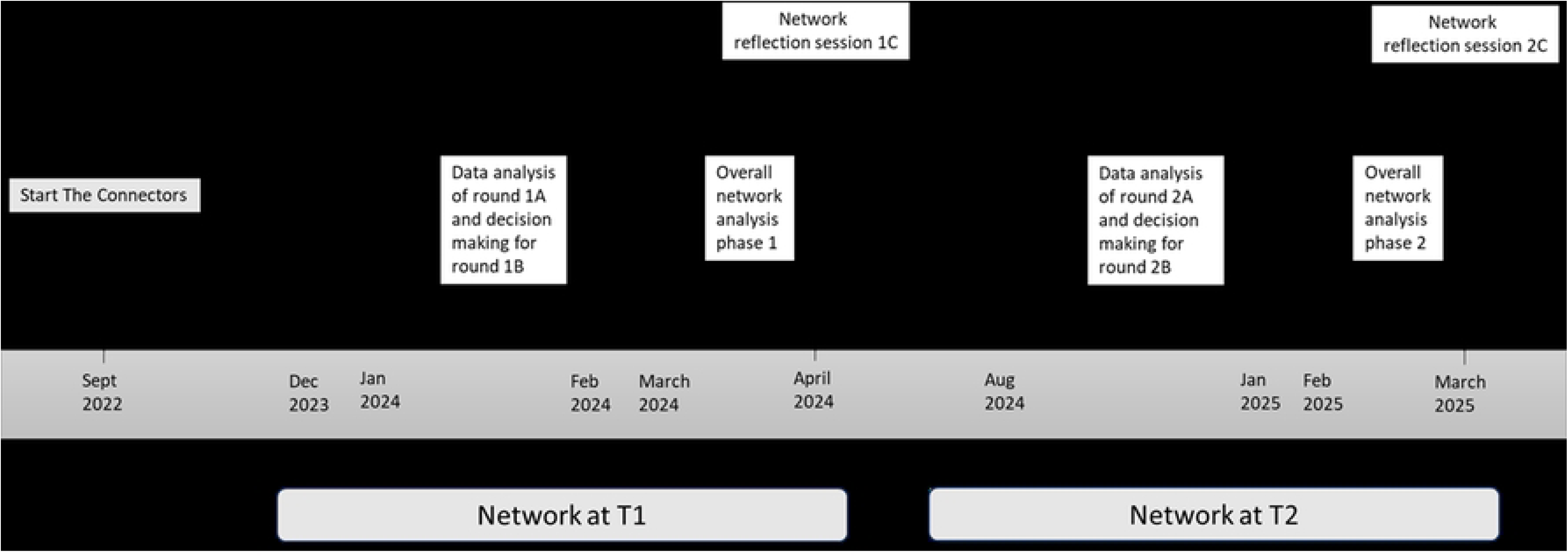
Timeline of the data collection.

We built on the interview instrument of Tijhuis (24) and De Jong et al (25) by drawing the network of the interviewees (The Connectors), while answering questions about the actors and relationships in the network (see S1 Appendix). The interview instrument for the first round of interviews with The Connectors consisted of 1) a name generator question to enumerate all individual actors with whom the respondent collaborates, 2) name interpreter questions to obtain information about the characteristics of those actors, and 3) a name interrelator question to identify mutual collaborations between actors.

Both in phase one and two, The Connectors were interviewed in round A. Then, by using a snowball sampling method, an extra round of interviews (round B) with the central actors mentioned in round A was conducted (26). The five most central actors in each individual network of the Connectors team members (based on relative degree), derived from the interview round A, were interviewed in round B. Accounting for overlap in central actors across the different networks of the team members, twelve interviewees were included in round 1B and seventeen interviewees in round 2B. No name interrelator questions were included in interview rounds B.

The purpose of the reflection sessions (1C and 2C) was to evaluate the networks compiled based on the interviews. We looked at both the individual networks of the team members (rounds 1A and 2A) and the distinct networks at T1 and T2. Three topics were discussed during these reflection sessions. First, we examined whether the networks were recognizable to team members (and why or why not). Next, we discussed the changes over time and finally, we discussed action points arising from the network visualization. For example, are there sectors or organizations that are less well represented in the network, but which do fit in with the questions received by The Connectors?

Other methodological choices were made to ensure the feasibility of the research, given time and resource constraints. A snowball sampling strategy was used to capture intra-team connections and to identify external actors mentioned as collaborators by their contacts (27, 28). This approach was particularly important because it allowed us to trace relational pathways beyond the initial team and to uncover actors who might otherwise remain invisible in formal organizational structures. At the same time, this method primarily reflects the more active and visible part of the network, and individuals not reached in subsequent rounds may remain excluded. As a result, weaker or peripheral relationships may be underrepresented. Previous research suggests, however, that the set of most central actors usually carries the greatest explanatory power for understanding collaboration processes and role shifts (21, 29). It is important to note that centrality does not only refer to actors with many ties, but also to bridging actors who, despite fewer connections, play a crucial role in linking otherwise disconnected groups. In the specific context of neighbourhood-based intersectoral collaboration, focusing on central actors is particularly meaningful: professionals often occupy visible positions across multiple domains, while bridging citizens are expected to maintain numerous ties in the neighbourhood to connect services and community needs. A comprehensive overview of all actors would have been preferable, but given the research context, concentrating on these central and bridging figures provides the most valid and contextually relevant insights within the available resources. Moreover, network reflection sessions with *The Connectors* confirmed the distinct networks, which strengthened the robustness of the findings beyond the structured interview data alone (30).

The first phase of data collection started in December 2023 and the second in August 2024. This time interval was chosen to align with the relatively calm periods in the team’s annual work cycle, which enabled participants to engage in in-depth interviews. The interval between the first and second phases was approximately one year. This is long enough to allow for potential shifts in collaboration patterns to emerge.

#### Ethics

This study was conducted in accordance with the Declaration of Helsinki. The Medical Ethics Committee of the Leiden University Medical Center has mandated the review of research not falling under the Medical Research Involving Human Subjects Act (nWMO) to individual nWMO committees. The relevant nWMO committee reviewed the proposal and issued a declaration of no objection (non-WMO approval code: 23-3042, approval date: 5 June 2023). The study was further approved by the Science Committee of the Public Health and Primary Care department at Leiden University Medical Centre (approval code: WSC-2023-02/PvN, approval date: 20 April 2023).

Informed consent was obtained from all interviewees involved in the study.

### Analytical strategy

We compiled a squared sociomatrix of all mentioned individuals after both rounds of each phase based on the name generator questions. This therefore includes both the individual networks of the team members resulting from interview round A and the data collected during interviewed round B. The answers to the name interpreter questions were merged into a standard actor-by-variable matrix. This enabled the construction of two undirected networks (T1 and T2) representing the evolving structure of intersectoral collaboration and citizen involvement. All ties were treated as undirected throughout the analysis, which means that the ties connect two actors without a direction of the relationship (31). This decision was based on the sampling procedure, and the design of the name generator questions (S1 Appendix). Not all actors were interviewed, and therefore reciprocity could not be systematically verified. We assumed that a reported tie indicates a relationship regardless of whether it was reciprocated, which is consistent with the interpretation of collaboration in this context (32). Consequently, degree measures were calculated without distinguishing between in-degree and out-degree.

The matrixes were uploaded to R for the visualization of the network and the corresponding analyses. The answer to the open question (S1 Appendix, nr. 11) was transcribed. Also, the notes of the reflection sessions were written out. All qualitative data were imported to ATLAS-ti 22. This data is used to qualitatively interpret the changes of the relationships in the network of The Connectors and will be discussed together with the quantitative analysis.

To analyse the networks, we applied a range of Social Network Analysis (SNA) metrics (see S2 appendix for a complete list of the metrics and the definitions) at both the network and actor level. At the network level we first assessed the overall size and growth of the network by calculating the number of actors and relationships in the network. To evaluate structural changes over time, we used the Jaccard index. This captures the extent of change in relationship patterns, including the emergence and disappearance of relationships (31). It is calculated as “The size of the intersection divided by the size of the union of the two networks” (33). The clustering coefficient was calculated to assess local cohesion (31).

At the actor level, we started with a description of the actors included. The degree centrality reflects the level of activity or embeddedness of an actor in the network (31). To capture changes in actor roles over time, we compared the centrality scores between T1 and T2, identifying shifts in prominence and influence. Particular attention was paid to actors representing the community voice, whose presence and centrality were tracked to evaluate their inclusion and evolving role within the network. These metrics allowed us to assess whether citizen actors became more integrated and influential over time.

## Results

### Descriptive statistics

In total, the first phase included four interviews in the first round and twelve interviews in the second round. All together at T1 these interviewees mentioned 102 different actors, originating from ten different sectors. The second phase consisted of seven interviews in the first round and seventeen in the second round. At T2, 215 different actors were mentioned by the interviewees, originating from fourteen different sectors. Table 1 provides descriptive statistics of the network. Over time, the network more than doubled regarding the actors and relationships. The average degree, or average number of actors one actor collaborates with, slightly increased from 3.67 to 4.40.

**Table 1.** Descriptive statistics of The Connectors’ undirected network at T1 and T2.

|  | **T1** | **T2** |
| --- | --- | --- |
| **Number of actors** | 102 | 215 |
| **Number of sectors** | 10 | 14 |
| **Number of relationships** | 187 | 473 |
| **Average degree** | 3.67 | 4.40 |
| **Clustering coefficient** | 0.67 | 0.62 |
| **Actor overlap** |  | 0.17 |
| **Jaccard index** |  | 0.76 |

The clustering coefficients based on undirected ties measure the extent to which an actor’s direct relationships also have relationships with each other (31). Actors with a degree above one (in other words actors with more than one relationship) had a clustering coefficient of 0.67 at T1 and 0.62 at T2. This indicates a high level of local cohesion among the more active actors. While the overall clustering slightly decreased, Figure 2 reveals that the central core of actors became more densely interconnected at T2, with a larger number of actors forming reciprocal collaborations.

**Figure 2.**
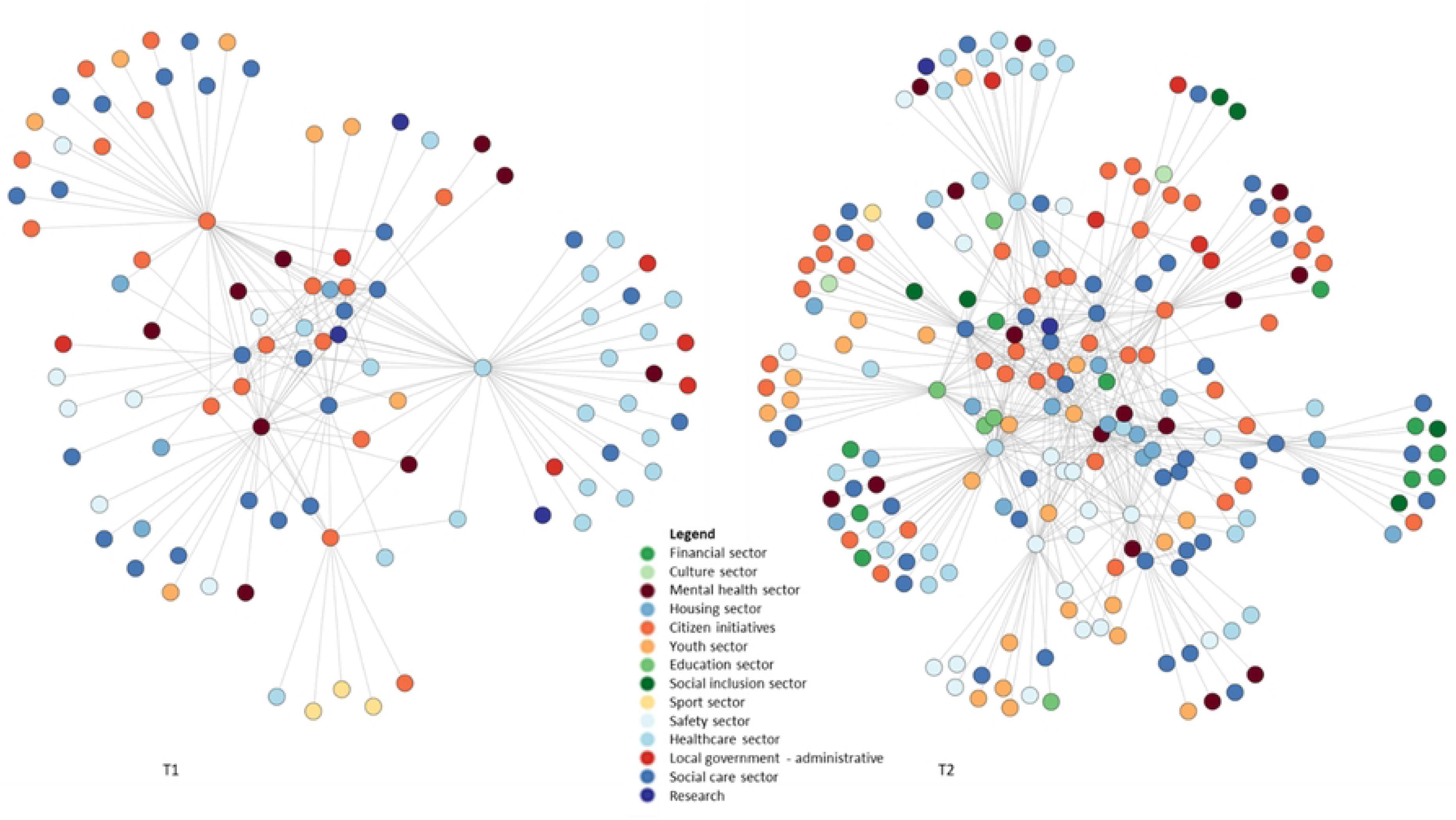
Network of The Connectors at T1 and T2.

At the same time, the network grew with 111% and became less centralized, as the degree centralization decreased from 0.399 at T1 to 0.241 at T2. This indicates that connections were distributed more evenly across actors. Moreover, new actors entered the network and formed local star-shaped hubs. These hubs connect peripheral actors who do not have a unmediated relationship with the team members, thereby extending the network’s reach and diversity. This structural evolution suggests a shift from a single highly centralized configuration (T1) toward a more distributed pattern of connectivity (T2). Consequently, the network may have become somewhat more resilient, as dependency on the initial core actors decreased. Improved network resilience means that the network can adapt better in the event of disruption or loss of actors/connections (34). Nevertheless, the persistence of hub structures implies that vulnerability remains: if one or more central connectors were to withdraw, parts of the network could still fragment (35).

Although the Jaccard index between T1 and T2 is relatively high (0.76), this measure refers only to the small subset of actors present at both time points (actor overlap = 0.17). Thus, the high Jaccard index reflects strong relationship retention within a narrow core rather than stability of the entire network. The network underwent substantial expansion. Many new actors appeared in T2, and a notable proportion of the relationships at T2 involve these new actors. Across sectors, the proportion of actors present at both measurement points was generally low, with the highest overlap observed among community initiatives (26.1%), see S3 appendix. Consequently, the stability in relationships was concentrated in a relatively small, stable core while the broader network experienced substantial turnover and growth.

A diverse group of actors is represented in the network. At T1 and T2, respectively 56.7% and 58.6% of the actors are women. At T1, fourteen actors were identified as having a dual role. This is divided into eleven actors primarily seen by interviewees as representatives of the community, while also performing professional tasks (and/or having a paid job) and three actors primarily seen by interviewees as professionals, while also representing the community. Five actors were considered community voice representatives without a dual role. At T2, twenty-eight actors were identified as having a dual role. This is divided into twenty-six actors primarily seen by interviewees as representatives of the community, while also performing professional tasks and two actors primarily seen by interviewees as professionals, while also representing the community. Thirteen actors were identified as community voice representatives without a dual role. These numbers indicate a significant overlap between professional and community-oriented roles in the network. In addition, there is a shift visible from actors with a dual role who are at T1 primarily seen by interviewees as representatives of the community voice to being seen primarily as professionals at T2. 71% of the actors with a dual role at T1 are still present at T2, but with a different focus of their dual role, according to the interviewees. Both at T1 and T2 19% of the total actors represent the community voice in some way. This means that the community voice representatives were involved right from the start of The Connectors but also took a strong position in the network during its further development. Reflection session A revealed that the citizen representative team member of The Connectors played an important role in establishing initial contact with other community voice representatives.

Table 2 and Figure 2 show that four new sectors have entered the network at T2 compared to T1: the financial sector (particularly debt assistance), culture, education, and participation. In addition, several existing sectors have expanded, e.g. organizations have joined from the social care sector, and more actors from the safety sector, such as neighbourhood police officers, have become involved. Figure 2 also illustrates that the network at T2 has developed a more centralized core with stronger interconnections between actors.

**Table 2.** Descriptive statistics of the actors included in The Connectors’ network at T1 and T2.

|  | T1 |  |  | T2 |  |  |
| --- | --- | --- | --- | --- | --- | --- |
|  | N (%) |  |  | N (%) |  |  |
| **Number of actors** | 102 |  |  | 215 |  |  |
| **Female** | 58 (56,7) |  |  | 126 (58,6) |  |  |
| **Role** |  |  |  |  |  |  |
| Professionals | 83 |  |  | 174 |  |  |
| Community voice representatives | 5 |  |  | 13 |  |  |
| Professionals with a dual role | 3 |  |  | 26 |  |  |
| Community voice representatives with a dual role | 11 |  |  | 2 |  |  |
| **Sector affiliation** | **N (%)** | **Core (%)** | **Periphery (%)** | **N (%)** | **Core (%)** | **Periphery (%)** |
| Social care | 25 (24.5) | 36 | 64 | 45 (20.9) | 40 | 60 |
| Citizen initiative | 18 (17.6) | 55.6 | 44.4 | 42 (19.5) | 35.7 | 64.3 |
| Healthcare | 20 (19.6) | 20 | 80 | 25 (11.6) | 24 | 76 |
| Youth | 7 (6.9) | 14.3 | 85.7 | 22 (10.2) | 45.5 | 54.5 |
| Local government - administrative | 6 (5.9) | 16.7 | 83.3 | 5 (2.3) | 60 | 40 |
| Safety | 7 (6.9) | 28.6 | 71.4 | 20 (9.3) | 55 | 45 |
| Mental health | 9 (8.8) | 55.6 | 44.4 | 15 (7.0) | 40 | 60 |
| Housing | 4 (3.9) | 75 | 25 | 15 (7.0) | 66.7 | 33.3 |
| Financial | - |  |  | 10 (4.7) | 20 | 80 |
| Social inclusion | - |  |  | 6 (2.8) | 33.3 | 66.7 |
| Education | - |  |  | 5 (2.3) | 60 | 40 |
| Research | 3 (2.9) | 33.3 | 66.7 | 2 (0.9) | 50 | 50 |
| Culture | - |  |  | 2 (0.9) | 0 | 100 |
| Sport | 3 (2.9) | 0 | 100 | 1 (0.5) | 0 | 100 |

During reflection session B all team members confirmed that the actors who are central at T2 play a crucial role for The Connectors, linking team members to a wide range of sectors. The network images of T1 and T2 were viewed by all team members, and everyone recognized the growth and indicated that the team’s daily work and associated contacts are well represented in the network. The intersectoral collaborations are seen as essential in nearly all cases handled by The Connectors, as they enable the provision of appropriate care and support to individuals in need. The social welfare domain is the largest sector in the network, both at T1 and T2. The demand for The Connectors began in the field of mental health, and the network shows that various sectors are involved, because The Connectors believe these sectors can contribute to solving the underlying problems of people with mental health issues. In addition, this network demonstrates the prevalence of citizen initiatives, as this is the second largest sector at T2. Citizen initiatives can, for example, be an initiative of women that organize activities for mothers in the neighbourhood or certain community centres. As can be seen in Figure 2 and Table 2, only the Sport and Culture sectors are positioned entirely in the periphery, all other sectors are more centrally embedded.

Representatives of all other sectors are located both closer to the core of the network and into the periphery. We operationalized periphery as actors with one or no connections in the network. Peripherality in the network is therefore not structurally associated with a specific sector but appears to be distributed across sectors.

### Dynamics of the network

Figure 3 visualizes the differences between the two networks. Figure 3 shows that there has mainly been an addition of actors to the network rather than a replacement of existing actors. The network of The Connectors is therefore a network that deepens and broadens without losing existing relationships too much. The disappearance or addition of actors and relationships from the network can be explained in part by actor turnover, but also in part by the changing demands of the network at T2, where other knowledge and skills are becoming more important. An example of this is the questions surrounding debt counselling or the number of cases involving children and therefore also affecting schools. The financial and education sector have been added to the network because of the questions that arose and relate to those sectors.

**Figure 3.**
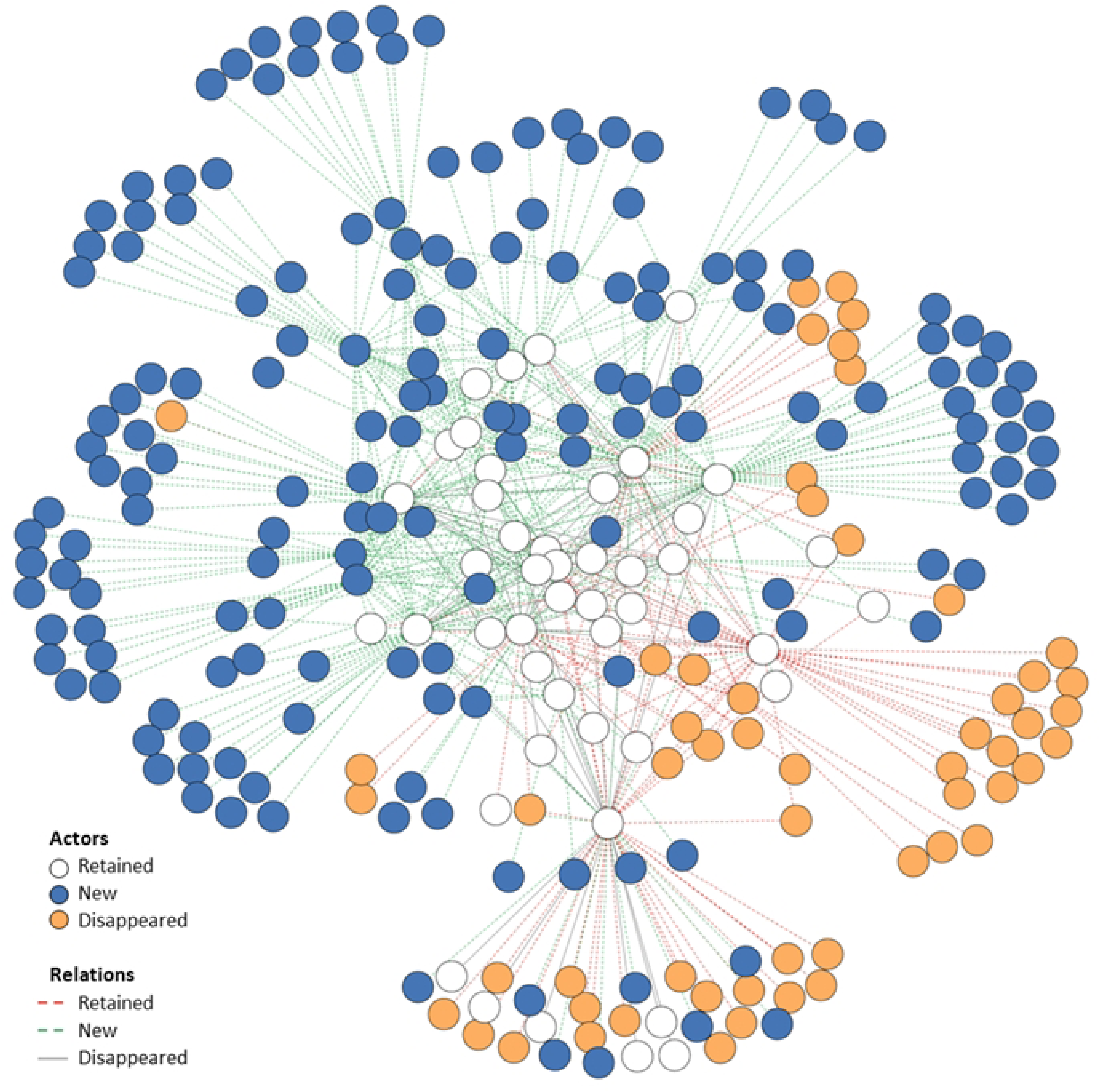
Network changes between T1 and T2.

When we zoom in on the actors that left the network after T1, based on the qualitative data, we conclude that approximately half of them can be attributed to personnel turnover. The other half of the actors that left the network turned out to be too far removed from The Connectors’ stricter goals developed over time, which meant that they no longer appeared in the day-to-day work of the core actors of the network. These changes were discussed in reflection session B, and the team members confirmed that the actors who had disappeared were no longer relevant and that contacts had faded over time. However, a bond had been established, so if the actors needed each other again at a later point in time, they would know how to reach out to each other.

The visualization of Figure 3 shows this described effect. For example, one team member in T1 and T2 (white actor in the upper part of the network, connected to both blue, purple and white actors) lost relations and added new relations to the network due to personnel turnover. In addition, several actors with whom this team member had a relationship came from the directors and managers of different organizations. The team member indicated in the interview that these policy actors were relevant in the initial phase of setting up the network and connecting organizations and sectors to the network, but in the T2 phase this proved to be less relevant in day-to-day activities.

These actors may be reconnected at a later stage if policy choices must be made, for example with regard to sustainable assurance issues. Another team member (white actor, left part of the network, who mainly has relations with blue and white actors) got a different role in the team and the network and realized parts of the peripheral actors did not add enough value to the network goals and lost contacts.

Between T1 and T2, the network expanded to include four additional sectors (financial, social inclusion, education and culture), as shown in the descriptive statistics. This growth was primarily driven by the requests The Connectors receive of citizens in need for help. The team responded to incoming requests for help with problems related to diverse problems by engaging the relevant sectors that could provide appropriate support. Furthermore, Reflection Session A revealed that a key care organization, despite its significant role in the neighbourhood for citizens, was not adequately represented in the network. As a result, one of the team members initiated contact, leading to the inclusion of this organization at T2.

At both measurement points T1 and T2, the majority of the collaborations occurred across sectoral boundaries. At T1, 75% of all relationships are between actors from different sectors, compared to 25% within-sector relationships. By T2, this proportion had increased slightly to 79% intersectoral relationships. This suggests that as the network expanded, new collaborations were not only confined to existing sectoral clusters but increasingly bridged different sectors. Although the proportion of intersectoral ties increased only modestly (from 75% to 79%), this change should be interpreted in the context of substantial network turnover and expansion between T1 and T2. Both at T1 and T2, most relationships are between actors of citizen initiatives and the social care sector.

The next most common intersectoral relationship (at T1 and T2) is between actors of mental health care and social care sectors. This pattern aligns with the qualitative findings in both reflection sessions that professionals and community voice representatives increasingly engaged beyond their traditional boundaries, which may enhance the network’s overall adaptability and responsiveness to complex mental health challenges.

### Actor turnover

The interviews and reflection sessions show that there was a difference between organizations in the network in how the transfer of personnel changes is handled by the actors. In the past year, several personnel changes have taken place in a number of organizations in the network of The Connectors. Several personnel changes have also taken place within The Connectors team. Interviewees indicated that a smooth transfer took place in one organization: the new actor was properly trained, and relevant knowledge was transferred, ensuring a smooth transfer. The collaboration between The Connectors and this organization continued only with a new actor as point of contact. This demonstrates that collaboration between The Connectors and the organization concerned is embedded within the organization and does not depend on a few individuals.

However, several interviewees indicated that this was not the case at another organization in the network with high staff turnover. Every time an actor leaves, contact must be re-established with this organization to rebuild the relationship from the start. During reflection session B, one of the team members stated: “This collaboration is necessary to help part of our target group, but it is difficult.” During the reflection session, The Connectors’ team leader identified the need to discuss with this organization how to ensure sustainable collaboration despite personnel changes. As a result, the team is committed to increasing the resilience of the network.

Figure 3 shows that in addition to the changes in actors in the network, new relationships have also emerged between existing actors. The team members got to know each other’s key contacts. This contributes to the network’s redundancy, which means that more than one path exists between two nodes in a network (31). Network redundancy is of added value when a team member is absent or may leave. It also enhances robustness by ensuring continuity of relationships and functions. Redundancy plays an enabling and reinforcing role in this network because multiple actors possess similar knowledge of the network through their shared contacts. This was confirmed in reflection session B in which team members indicated that cooperation with the central core has strengthened and the increased redundancy is appreciated, despite the added potential for red tape and the additional coordination required. There is mutual trust and they know what kind of questions and support they can turn to each other for. One of the collaboration partners interviewed described collaboration with The Connectors as follows: “By also taking a moment to ask the other person how things are going when you don’t have a request for help yourself, the cooperation strengthens.”

### Community voice and its actors

In the neighbourhood and in the network of The Connectors, there are citizens who play an active role and are indicated as community voice representatives (see Table 2). Some citizens do this in the form of maintaining many social contacts in the neighbourhood and signalling when someone is in need for care. Others have a volunteer role or have progressed from the volunteer role into a professional in the neighbourhood such as coordinator of a community centre. Because The Connectors operate in the neighbourhood and see an important role for community voice in their work, we also asked whether one sees an actor from their network as part of the representatives of the community voice or professionals of the network in both the round 1 and round 2 interviews of the network data collection (phase 1 and 2). When we zoom into the five most central actors of each team member (derived from the first and third round of interviews), respectively 38,5% and 45% of the actors is indicated as representatives of the community voice whether with a dual role. When we look at the proportion of actors representing the community voice in the entire network, this remains constant over time at 18.6%. This includes the sum of the roles of community voice representatives, community voice representatives with a dual role, and professionals with a dual role in Table 2. Thus, the community voice indeed plays an important and stable role by being present in the network of The Connectors both at T1 and T2.

It is noteworthy that all citizens representing the community voice who are present in the network at T1 are also active at T2. However, at T2 82% of the actors with a double role are now seen as professional as superior role in the network instead of community voice representatives. Their position in the network developed over time, without a change in the actual tasks of these actors in the network. We observed that actors representing the community voice have expanded their collaborations with other professionals in addition to the collaborations with The Connectors at T2. Earlier, these citizens indicated that collaborations with professionals were often difficult and not reciprocal. At T2, however, all actors were resolute in labelling all actors, including the actors representing the community voice, as professionals. Those community voice representatives seem to be taken more seriously in the collaboration between professionals and citizen.

The evolution from citizen representing the community voice to professional also involves complicated situations. These situations are influenced by the dual roles these actors have in the network. They are both citizen and professional in the same neighbourhood. This may lead to tensions in the relationships in the network. We observed three types of mechanisms underlying the tensions arising from the dual roles of actors during data collection. First, we identified the pressure to be a professional first, and only then a citizen. One of the actors whose role changed from citizen representing the community voice to professional, indicated that she sometimes skips citizen gatherings she used to frequent, even though she is a citizen of the neighbourhood herself as well. She experienced that since she has the professional role in the neighbourhood, both other professionals and citizens find it difficult to see her outside that role, even in informal settings. She indicated: “Sometimes it feels like as soon as you step out the door you are the professional, regardless of whether it is during your working hours or not.” The second mechanism regards a strain on personal relations in the network. An example of this mechanism observed in the data collection illustrated how tensions between neighbours can arise. A citizen asked for help from a neighbour who is not only a citizen but also a professional working in the neighbourhood. When this help cannot be provided (as desired), this jeopardizes the personal relationship between the neighbours. Finally, we identified a tendency toward conflicts in the network. Tensions emerged when a professional dispute arose involving an actor who holds a dual role in the neighbourhood, both as a professional and as a citizen. The conflict was known to other actors in the network. The actor with the dual role had to continue to interact with the same professionals in everyday life, not only in their professional capacity but also as a fellow citizen. The dual role complicated the dynamics of the conflict which had implications for the actor’s relationships within the network. The professional engagements were affected by the continued personal interactions in the neighbourhood setting.

The situations as described show that the actors who develop from a citizen representing the community voice to a professional in the network will also have to develop skills to deal with the dual role they fulfil in the neighbourhood from now on. It requires more guidance and awareness of the dual roles among the other actors in the network. As someone pointed out in an interview, reflecting on one of the above complicated situations that occurred for actors with a dual role: “it is a curse and a blessing at the same time to live and work in the same neighbourhood”.

## Discussion

This study examined the development of a local intersectoral network in a low SES neighbourhood, with a particular focus on the evolving roles of representatives of the community voice and professionals in addressing (mental) health needs. Building on the aim to understand how community voices are integrated into intersectoral collaboration, our longitudinal social network analysis revealed that The Connectors team successfully expanded the network over time, both in size and diversity. By T2, the network had grown to include a broader range of sectors and actors, while largely maintaining existing relationships. This expansion was accompanied by a growth in intersectoral relationships at T2, suggesting that The Connectors fulfil their intended role as boundary spanners (36). These developments reflect a maturing network structure that supports deeper collaboration between professionals and community representatives.

We observed three phenomena in the development of the network in the low SES neighbourhood, which shed new light on intersectoral collaboration including the community voice. First, with the inclusion of the community voices, citizens became part of the network. The role of citizens representing the community voice developed over time to professionals as well, as observed in the network at T2 compared to T1. This dual role as both community voice representatives and professionals can be empowering, offering pathways to personal development and career progression. Our findings confirm the existing literature that shows that dual roles provide among others opportunities for reciprocity, increased perceptions of trust and community accountability (37). However, it also introduces complexity. As described in the results, actors with a dual role may face tensions or even conflicts of interest, that require an ethical decision (38, 39). We add three underlying mechanisms for these tensions to the existing literature, namely 1. the pressure to be a professional first, and only then a citizen, 2. a strain on personal relations in the network and 3. a tendency toward conflict within the network.

From a network perspective, it is crucial that these actors remain connected to the broader community to continue representing the community voice authentically, because there is a risk that those actors align more with institutional logics and lose representativeness and downward accountability to their community (40, 41). If this connection weakens, the network should proactively identify and support new representatives to maintain inclusivity and responsiveness.

Second, over time, the request of citizens in need for help drove network expansion and diversity. The data showed that this network expansion did not confine to existing sectoral clusters, but bridged different sectors, namely additional sectors were added. The Connectors are therefore actively working to translate citizens’ needs into the professional network in order to improve the assistance offered. In addition, they contribute to bridging sectors, and we can conclude that cross-sectoral cooperation has grown over time.

The third phenomena is that structured reflection sessions on the network can serve as an effective tool for onboarding new team members. The network experienced high personnel turnover, a common challenge in community-based and interdisciplinary initiatives. By visualizing the network and discussing its structure and dynamics, new employees gained insight into existing relationships, collaboration patterns, and strategic positions within the network. This supports not only knowledge transfer and retention, but also team cohesion and strategic alignment. Previous studies have highlighted the value of SNA for organizational learning and network development (42). Our study contributes to the existing literature by demonstrating its utility for onboarding and continuity in dynamic team environments (12). Moreover, the periodic use of SNA combined with the structured reflection sessions enabled The Connectors to reflect on their own practices and actively steer collaboration efforts.

This study has two main limitations. First, the longitudinal network analysis was conducted over a relatively short timeframe of three years from the start of The Connectors. Although a longer follow-up period would provide deeper insights into sustained network dynamics, the observed changes already contribute to the literature on emerging intersectoral collaboration. Second, the structure of the network was partially shaped by the design of the data collection, which began with interviews in a single team and expanded via snowball sampling. As a result, not all actors had equal opportunity to nominate their collaborations, potentially underrepresenting certain relationships.

Beyond these methodological considerations, a substantive limitation concerns the assumption that collaboration itself leads to improved outcomes in mental health care. Whether such outcomes are indeed achieved, and how factors such as dual professional roles influence effectiveness, remains an open question. Future research should therefore examine the concrete impact of intersectoral collaboration including the community voice on client well-being.

## Conclusion

We explored how networks emerge and evolve over time and focussed on the development of the roles of citizens in the network and diverse sectoral involvement within a local network in a neighbourhood with low socioeconomic scores. Particular attention is paid to the dynamics between professionals and citizens who represent the community voice, and how their collaboration deepens and transforms as the network matures. This study shows that the community voice is crucial in the network, among others as a gateway to the rest of the neighbourhood’s residents and in order to be able to offer the right help in the neighbourhood. It is beneficial to include a community voice representative in the team or the nascent network so that the team, together with the community voice representative, can then perform a boundary spanning role between professionals and community voice representatives.

Strengthening intersectoral networks that include both professionals and community voice representative requires time and flexibility. Our findings highlight the importance of fostering both professional relationships and relationships with community voice representatives and acknowledge predictable change in the role of the latter. Moreover, the importance of creating space for collaboration across sectors is central. To enable such network development, policies must allow room beyond standard protocols and organizational silos, as well as provide sufficient time for relationships and structures to mature. Although network building is a gradual and complex process, once established, these networks can play a pivotal role in delivering integrated and responsive care.

## Data Availability Statement

The qualitative and quantitative datasets generated and analyzed during this study contain potentially identifying information due to the small participant group and the presence of (indirect) identifiers. In accordance with privacy and ethical requirements, the full datasets cannot be made publicly available. De-identified quantitative data (with selected variables removed to reduce re-identification risk) may be shared by the authors upon reasonable request, provided that such sharing complies with applicable privacy safeguards.

